# A survey and review of eye-tracking in clinical practice

**DOI:** 10.64898/2026.09.07.26361936

**Authors:** Fiona B. Mulvey, Onyekachukwu Mary-Anne Amiebenomo, Denize Atan, Siyuan Chen, Amanda Douglass, Matt J. Dunn, Daniel Goldstone, Rasha Sameer Moustafa, Frederic Shic, Gemma Arblaster

## Abstract

**Purpose:** The aim of this study was to evaluate how eye-trackers are currently used in clinical settings and to explore whether guidelines are perceived to be needed to support their use in a clinical context.

**Methods:** A survey was developed and distributed to clinical eye-tracker users, manufacturers, professional bodies and other key stakeholders. The survey explored current practices, perceived barriers to clinical eye-tracking, guideline availability, and the perceived need for guidelines to support clinical practice.

**Results:** Eighty responses to at least one of the survey questions were received. Of these, three were excluded from some analyses due to having no clinical involvement in eye-tracking (their involvement was in research only). Eye-tracking was performed by ophthalmologists, orthoptists, neurologists, optometrists, allied scientists and audiologists. Whilst most respondents were assessing eye movements daily, eye-tracking was mostly performed either rarely (less than 10%) or moderately often (10-60%) in their practice. Most clinical respondents (76%) reported they both performed eye-tracking and interpreted the results. The most commonly identified barriers to clinical eye-tracking were eye-trackers being considered primarily research tools, considered unsuitable for performing the assessments, and lack of access to an eye-tracker. Most respondents (71%) were unaware of any existing guidelines for clinical eye-tracking. Where guidelines were reported to exist, they were described as somewhat sufficient to completely insufficient. Most respondents (93%) supported the development of guidelines for clinical eye-tracking. A small number of respondents (7%) reported concerns about introducing clinical eye-tracking guidelines.

**Conclusion:** A broad range of clinicians, professionals and other stakeholders involved in clinical eye-tracking responded to this survey. Their responses suggest a general consensus of a need for clinical eye-tracking guidelines. Whilst guidelines may address some of the reported barriers to eye-tracking clinically, and support benchmarking and standardisation across a broad range of clinical disciplines, concerns around a perceived threat to autonomous clinical practice should also be considered.

## Introduction

Recent decades have seen eye-tracking technologies extend from research into clinical practice. Eye movement measures obtained from eye-tracking systems provide an accessible and quantifiable index of neural integrity, reflecting the integrated function of the oculomotor and vestibular systems and their underlying brainstem, cerebellar, and cortical networks [6, 42]. Eye movement measures are used in the diagnosis and monitoring of ophthalmic, neurological, and psychiatric disorders [19]. The transition from electro-oculography (EOG) and magnetic scleral search coil (SSC) systems to infrared video-based oculography (VOG) during the early 2000s enabled precise, non-invasive measurements suitable for clinical contexts [29, 31, 48], and eye-tracking became a practical clinical tool. This translational shift is described in a review by Larrazabal et al. [41]. Yet despite technological progress, implementation remains inconsistent, and the absence of harmonised standards for acquisition, calibration, and interpretation continues to limit comparability across sites, devices, and datasets [33, 86], and therefore also across patient visits and specialities.

Eye movements are unique among physiological signals because they are both easily observable and deeply informative about distributed neural systems. From the pioneering work of Robinson [64], Bahill, Clark & Stark [4], and Collewijn [20], who used substantially different tracking methods and hardware, to modern infrared video-based systems, the field has refined its capacity to capture ocular motion as a direct biomarker of neurological function. More recent analyses have revisited classic saccade metrics, comparing them to current methods for computational precision [26]. An increasing diversity of recording systems intended for a wide range of clinical, research, and consumer usage scenarios have become commercially available. Early EOG techniques [10] provided low-cost, long-duration measures, while magnetic SSC systems [36] established the benchmark for accuracy. The development of VOG has since produced systems varying in accuracy, precision, comfort, and accessibility, with several systems enabling quantitative eye-movement assessment in outpatient and bedside settings. These variations in tracking methods and hardware may significantly affect eye movement measurements which may subsequently affect clinical decisions.

### Expansion of eye-tracking across clinical domains

Applications of eye-tracking have proliferated across medicine and psychology. In vestibular and balance disorders, the movements of the eye remain the primary indicator of labyrinthine asymmetry. Tests such as caloric irrigation, the head impulse test, and positional manoeuvres that exploit the vestibuloocular reflex (VOR) are performed in order to localise peripheral and central lesions [18, 27, 29, 48]. Structured diagnostic criteria from the Bárány Society now define presbyvestibulopathy [2], bilateral vestibulopathy [71, 73], and acute unilateral vestibulopathy [72] with consistent clinical taxonomy. Quantitative biomarkers have also been proposed for vestibular migraine [3] and comparative caloric versus electrical vestibular stimulation [49]. Video head impulse testing (vHIT) and VOG recordings allow objective quantification across age groups, including children [79], while recent normative studies have refined thresholds for binocular and torsional responses [69, 70].

Eye-tracking also provides diagnostically relevant measures across a range of neurological disorders and demyelinating conditions [25], Alzheimer’s disease and dementias [83], and psychiatric disorders [77]. In Parkinson’s disease (PD), saccadic slowing, reduced blink rate, and smooth pursuit deficits are established biomarkers [30, 80]. Modern VOG and artificial-intelligence methods now detect subclinical patterns in microsaccades and vergence [57, 87], providing non-invasive biomarkers of dopaminergic dysfunction. Disease-specific oculomotor phenotypes have been reported in progressive supranuclear palsy (PSP), where early slowing of vertical saccades and frequent square-wave jerks are characteristic oculomotor features that help differentiate PSP from idiopathic PD [22, 60, 88]. The differentiation of movement disorders by oculomotor profile exemplifies the broader potential of eye-tracking to support early diagnosis and subtype classification, extending far beyond what is directly observable in clinical examination [38, 44].

Wearable, head-mounted eye-tracking systems have been used to confirm distinctions between PD and PSP [28, 32]. In multiple sclerosis (MS), internuclear ophthalmoplegia (INO) is readily detected by saccadic disconjugacy measured during binocular viewing [8] and additional oculomotor abnormalities have been associated with broader cognitive dysfunction [23, 56]. Studies comparing VOG with MRI, in INO due to MS, have demonstrated that functional abnormalities may be detectable even when corresponding lesions are not evident on MRI [58].

Cognitive and traumatic disorders are an expanding application domain for clinical eye-tracking. In mild cognitive impairment, dementia, and Alzheimer’s disease, eye-movement measures show characteristic slowing, fixation instability, and biases in visual attention that correlate with cognitive decline [25, 45, 47, 62, 83]. Eye movement parameters and pupillary indices have also been shown to be robust biomarkers of recovery from traumatic brain injury (TBI) [16, 50, 51, 75, 81]. Recent studies outline diagnostic strategies for vision and oculomotor disorders following concussion [1, 65] and review diagnostic strategies for TBI [15], including large-scale classification and device-comparison [40].

Eye movements have been established as a sensitive measure of function in other conditions such as disorders of the neuromuscular junction [17, 82], and amblyopia [66, 74], as well as in the assessment of gaze-evoked nystagmus [5]. Eye movements have also demonstrated value as biomarkers in a diverse range of conditions, including neurodevelopmental and psychiatric disorders such as autism spectrum disorder (ASD) [39, 67, 77], attention-deficit/hyperactivity disorder (ADHD) [61, 77], schizophrenia [43, 53], anxiety disorder [46] and anorexia nervosa [59]. Across these different domains and clinical conditions, eye-tracking provides a convenient, repeatable, and multidimensional signal linking neural and cognitive health and function with non-invasively measurable behaviour.

### Measurement methods and data quality

The clinical utility of eye-tracking depends fundamentally on data quality and on the clinician’s ease of use under time pressures of clinical practice. Spatial and temporal precision, sampling rate, calibration, and noise handling can dramatically affect derived metrics, as consistently emphasised in methodological literature [33, 52, 63, 89]. Independent assessments of commercial VOG and video-nystagmography (VNG) systems have demonstrated variable precision across devices and contexts [24], while practical frameworks for recording and interpretation have been suggested for specific tests or procedures only [37]. Advances in noise modeling [85, 86] offer means of improving reproducibility across hardware, while population-based studies have shown that factors such as ethnicity and experimental design influence calibration accuracy [9, 33]. Clinical users face an additional challenge: patients may present with reduced visual acuity, limited fixation, ptosis, strabismus, limited eye movements, reduced cooperation or cognitive impairment, all of which can degrade data quality and diagnostic power unless systems incorporate robust validation and recovery routines [21].

Since many diagnostic benchmarks were derived from legacy recording systems that differ substantially from modern hardware, harmonised standards would ideally accommodate both contemporary and legacy technologies. Comprehensive procedural discussions such as the Oxford Textbook of Vertigo and Imbalance [14] and The Neurology of Eye Movements [42] address many of these practical aspects, but do not describe quantitative standards for eye data acquisition and analysis.

### Goals of the present study

Eye-tracking is now at a crossroads between research innovation and clinical application. Evidence is widespread for its utility in diagnosing, differentiating, and monitoring a wide range of neurological, vestibular, ocular and cognitive disorders. Yet, barriers remain to its reliable and convenient integration into routine clinical practice. Across the literature, calls for standardisation—spanning data quality, calibration, reporting, and interpretation—have grown louder over time [19, 33, 52, 86]. However, most initiatives to date have been researcher-driven rather than clinician-led.

Procedural guidance has advanced unevenly and is often fragmented between diagnostic definitions, educational materials, and manufacturer documentation, yet none offer a unified framework for clinical practice with support for data quality considerations in recording and clinical interpretation.

The present study aims to characterise the current landscape of clinical eye-tracking through a survey of clinicians, allied health professionals, or others who record or analyse eye-movement data in a clinical setting, and manufacturers of eye-tracker technology. By documenting how, when, and why eye-trackers are used across clinical settings and specialities, and by identifying clinicians’ perceived barriers to wider adoption, this work aims to provide a practitioner-oriented perspective for future consensus guidelines. The goal is to support and motivate the development of practical recommendations that respect scientific rigour, diagnostic precision, and the realities of time and training in clinical practice.

To this end, the International Society for Clinical Eye-Tracking (ISCET) was established in 2023 to promote international collaboration, knowledge exchange, and the development of open standards for clinical applications of eye-tracking [54, 55].

The survey was designed to answer the following research questions, specific to clinical use and applications of eye-tracking in human health:

#### How are eye-trackers used clinically?

- Who (or which specialisation) performs the eye-tracking, data analysis and interpretation?
- How frequently is eye-tracking performed?
- Which systems are used?
- Which measures or data are used, and by whom?
- How is data processed, analysed and interpreted?
- Are there limitations to, barriers or concerns about eye-tracking at present?

#### What guidelines, if any, are used for eye-tracking clinically?

- Where they exist, is there a desire or need to change existing guidelines?
- Where they don’t exist, is there a desire or need to develop guidelines?

## Methods

The ISCET survey working group developed the survey. It was written in English, piloted internally, and refined through feedback from the working group. The survey instrument was created in Qualtrics (Qualtrics, Provo, UT, USA). The investigation was carried out in accordance with the Declaration of Helsinki. Following ethical approval (University of Sheffield UoS064587) the survey was disseminated internationally to maximise diversity of responses. The survey was open to clinicians, professional bodies, manufacturers, and stakeholders across the clinical eye-tracking community. It was distributed electronically to 106 organisations identified as being relevant stakeholders by the working group and falling into the following categories:

### Clinical organisations focused on

**Clinical organisations focused on:**
|  |  |
| --- | --- |
| Audiology | Ophthalmology |
| Concussion | Optometry |
| Dizziness/balance | Orthoptics |
| Eye movement disorders | Otolaryngology |
| Neuro-ophthalmology | Otology |
| Neurodiversity & reading disabilities | Vision |
| Neurology/neuroscience | Visual electrophysiology |
| Neuropsychology |  |

### Eye-tracker manufacturers

Eye-tracking software developers (with potential clinical uses)
Regulatory bodies
Funding agencies

For the full list of organisations contacted, see Online Resource 1.

The survey was open between October 2024 and September 2025. No responses were received after March 2025. The survey questions are available in Online Resource 2. Prior to completing the survey, participants were presented with study information and gave their informed consent to participate. Data handling of the survey results is described in Online Resource 3.

If respondents had more than one applicable involvement with clinical eye-tracking (question 4), they were asked to select the option that best described their involvement and complete the survey in that capacity. Where respondents reported more than one speciality, both were retained except when calculating response rates. In Figures 1, 2 and 6 such respondents are counted once, under the speciality shown. Respondents are included only where they answered the question.

**Fig. 1.**
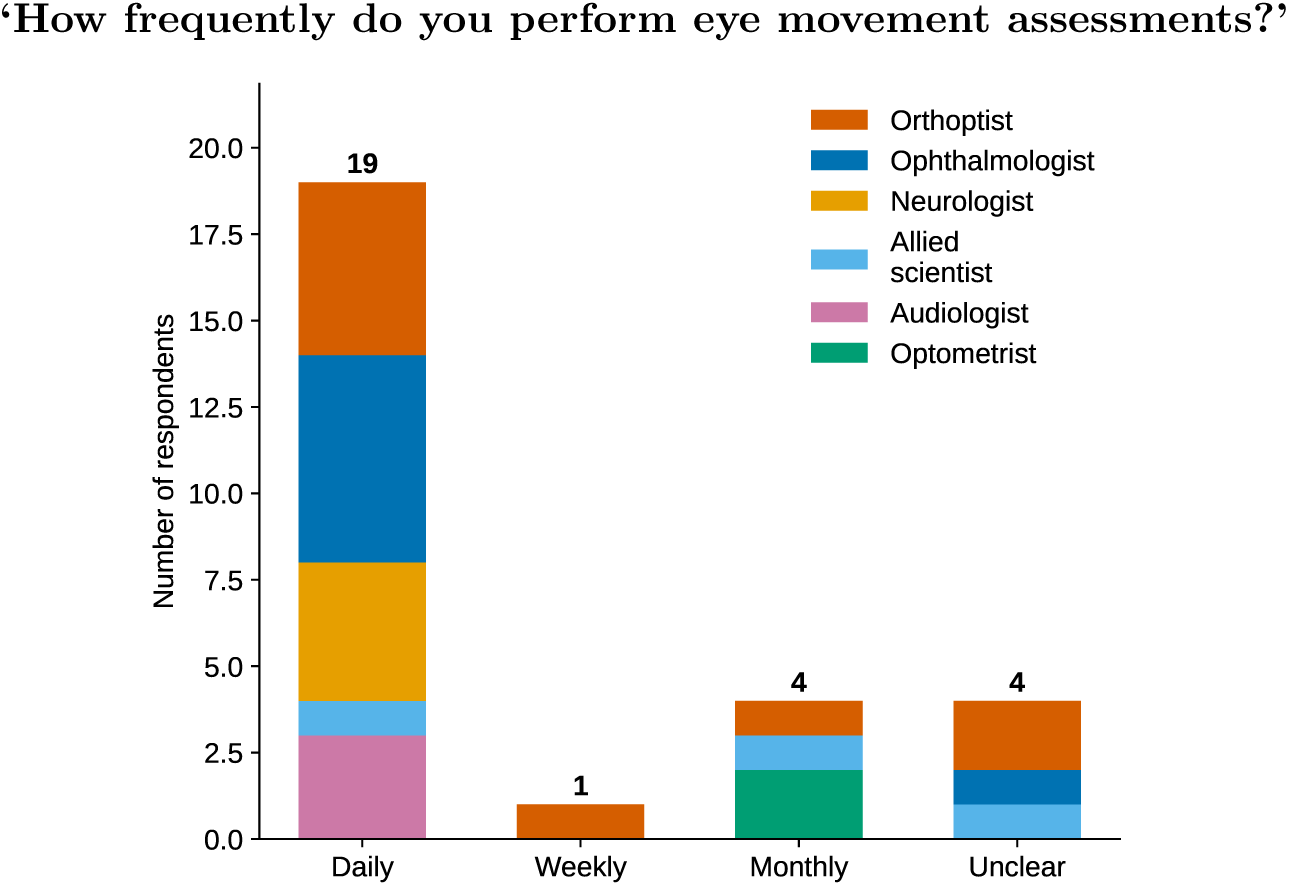
Session frequency of assessment of eye movements, stratified by clinical speciality. If clinicians did not specify a speciality, they are not included in this plot. One clinician specified two specialities (in ‘monthly’), and so are counted twice here. Some clinicians did not report assessment frequency and so do not appear in this figure.

**Fig. 2.**
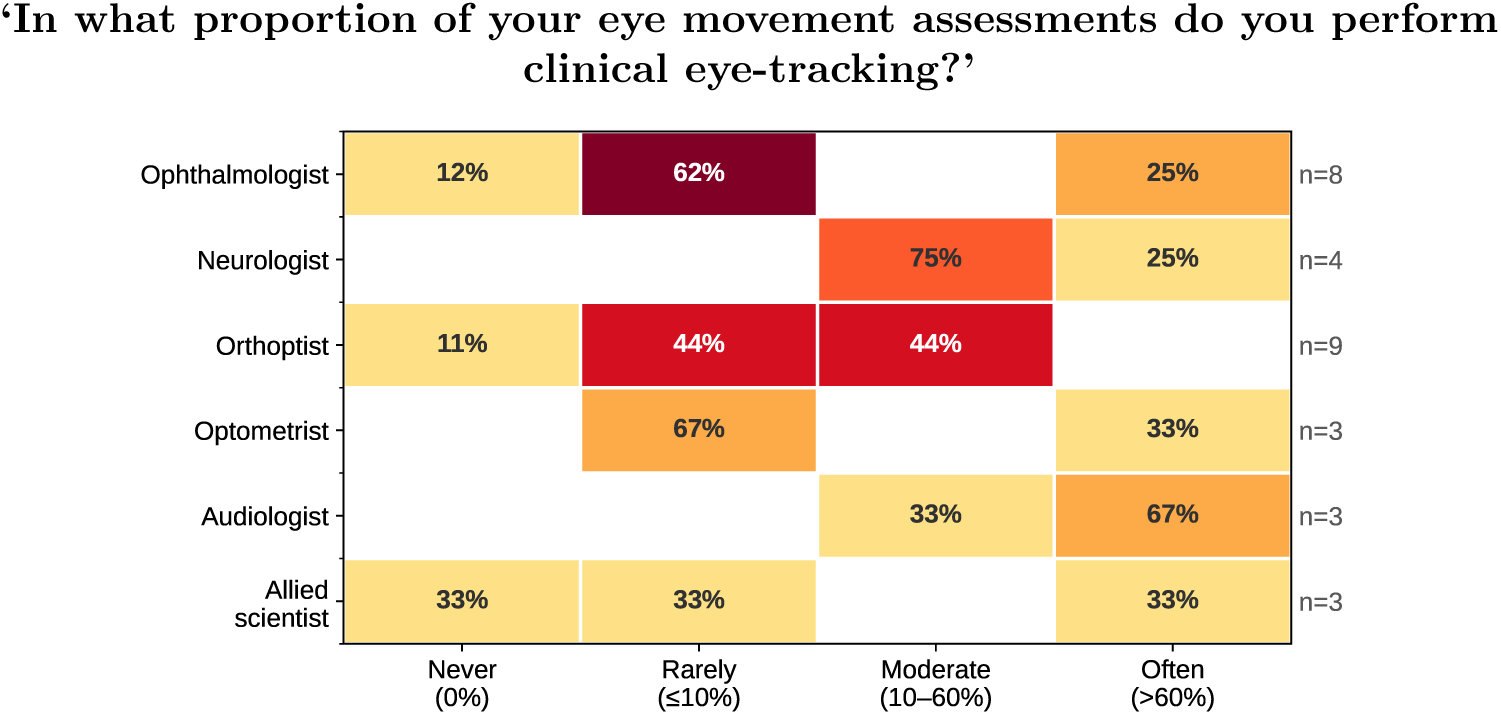
Proportion of eye movement assessments where eye-tracking technology is used, stratified by clinical speciality. If clinicians did not specify a speciality, they are not included in this plot. One clinician (in ‘rarely’) specified two specialities, and so is reported twice here

## Results

### Participants

One hundred and twenty-three identities were logged as opening the survey, and responding to at least one consent statement (there were seven statements related to consent for every respondent, see supplementary information ESM 2). Eighty participants responded to at least one survey question. Three responses from researchers who did not engage with clinical eye-tracking were excluded from some analyses. A further 43 read and reported they understood some or all of the participant information but did not participate in the survey: of these 43, 19 gave consent to all statements to participate, but gave no answers to any of the survey questions. The remaining 24 of 43 gave consent to some statements only, and they did not progress to the survey questions. See Table 1 for a full breakdown of participant response rates. Region or geographical information was not collected, as this would be potentially identifiable.

**Table 1.**
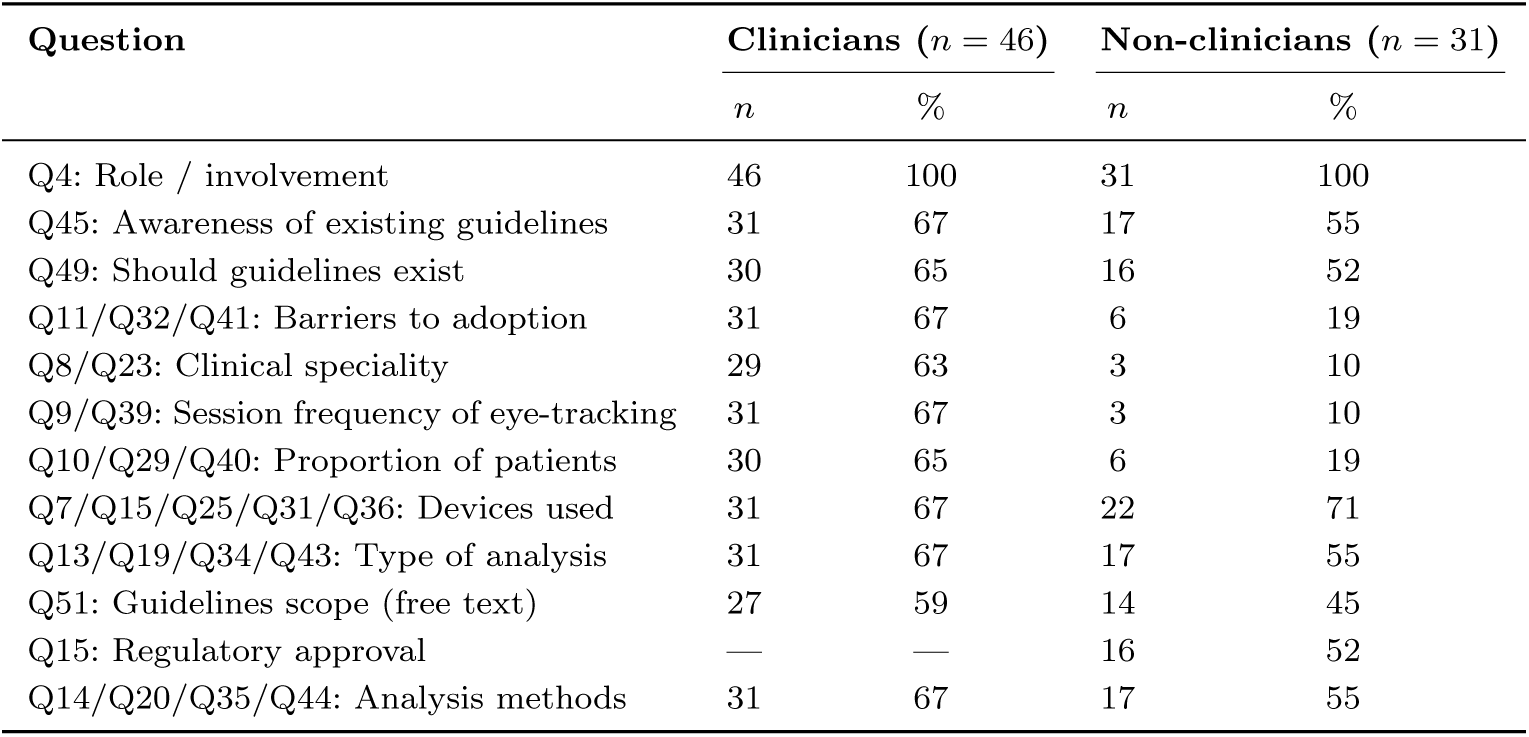
Response rates by question and respondent type. Clinicians are defined as respondents who perform or interpret clinical eye-tracking (Q4; *n* = 46). Non-clinicians are all remaining respondents who answered Q4 (*n* = 31), comprising manufacturers (*n* = 19), professional body representatives (*n* = 8), and those who selected ‘Other’ in Q4 (*n* = 4). A further 43 respondents did not answer Q4 or any subsequent question and are excluded. Percentages are calculated from the respective group totals. Q15 was a manufacturer-only question and is not applicable to clinicians.

### Current use of eye-tracking

There were 46 respondents who reported they perform clinical eye-tracking and/or interpret the results during their clinical duties (n=43), or described their role as ‘other’ but reported they were from a clinical institution (n=3). Twenty-seven of those 46 respondents gave their clinical speciality and described how frequently they assessed eye movements as part of their clinical role. Eye movement assessments were most commonly performed daily (see Figure 1).

The proportion of eye movement assessments where eye-tracking technology was used was most frequently reported to be *≤* 10% by 38% of respondents, although 22% reported using eye-tracking technology in over 60% of their eye movement assessments. Since the survey was likely completed by individuals with a pre-existing interest in clinical eye-tracking, it is not surprising that many respondents already used eye-tracking in their practice. However, the breakdown by clinical discipline is of interest (shown in Figure 2).

Most clinicians reported that they both perform eye-tracking and interpret the results (see Figure 3). Of the 48 respondents who described how results were interpreted and/or reported (31 clinicians, 17 non-clinicians), 44% (21/48) reported this was done both quantitatively and qualitatively, 29% (14/48) interpreted and/or reported only qualitatively, 21% (10/48) interpreted and/or reported only quantitatively, and 6% (3/48, all non-clinicians) were unsure what interpretation and/or reporting was performed, see Figure 4. Forty-eight respondents (31 clinicians and 17 non-clinicians) reported how eye-tracking results were analysed. Two selected ‘other’ but gave no further detail, 32 gave a single response and 14 gave multiple responses, giving a total of 65 responses. Analysis was with commercially available software (45%, 29/65), using own software (31%, 20/65), using open source code (12%, 8/65), no analysis performed (6%, 4/65) and unsure how the analysis was performed (6%, 4/65). The most popular eye-tracker named amongst respondents was the EyeLink from SR Research (27% – see Figure 5), although it is worth noting there are multiple devices in the EyeLink series, and several respondents reported ‘EyeLink’ without specifying which device is used.

**Fig. 3.**
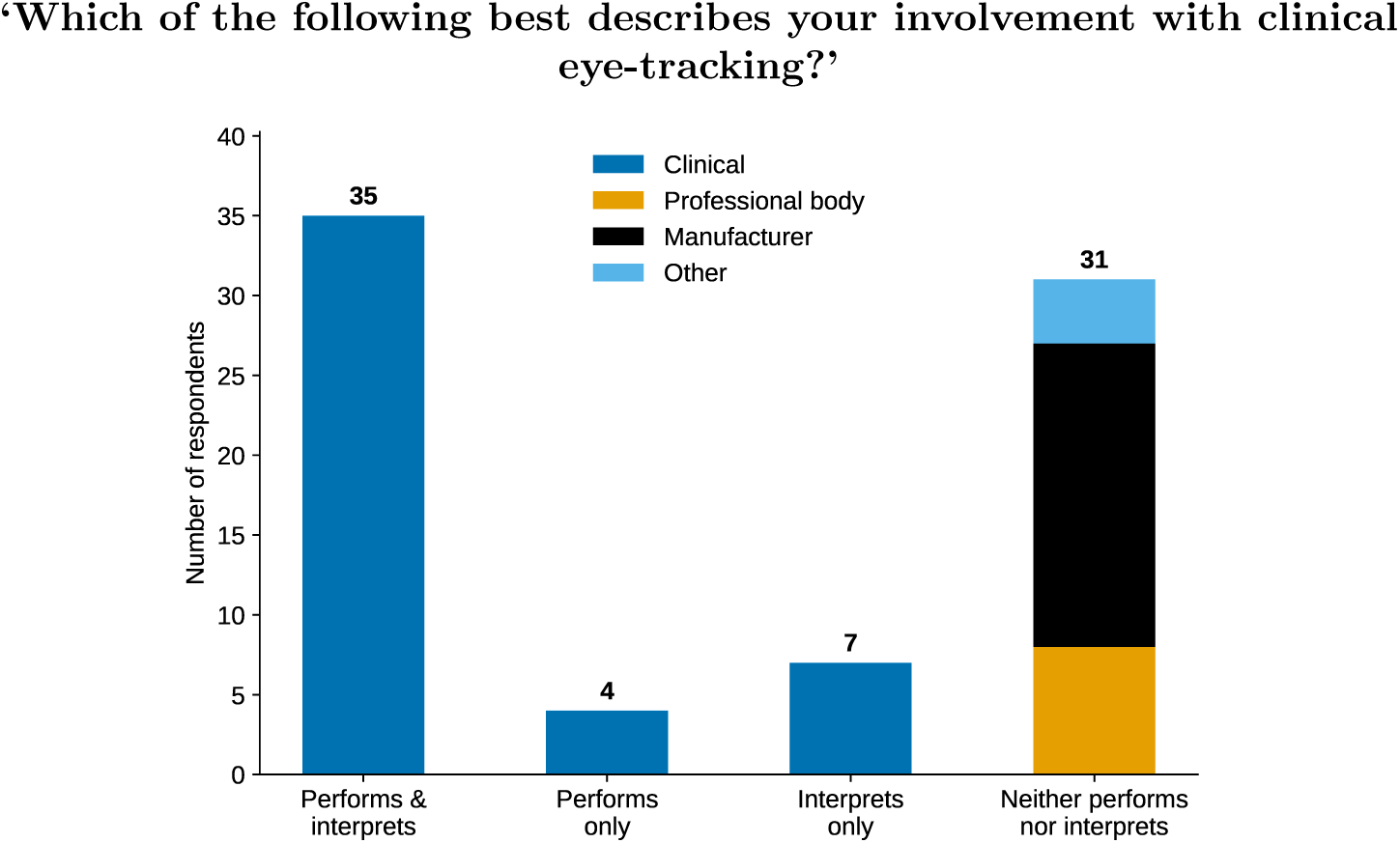
Respondent involvement with clinical eye-tracking by role, where ‘performs’ means records eye-tracking data

**Fig. 4.**
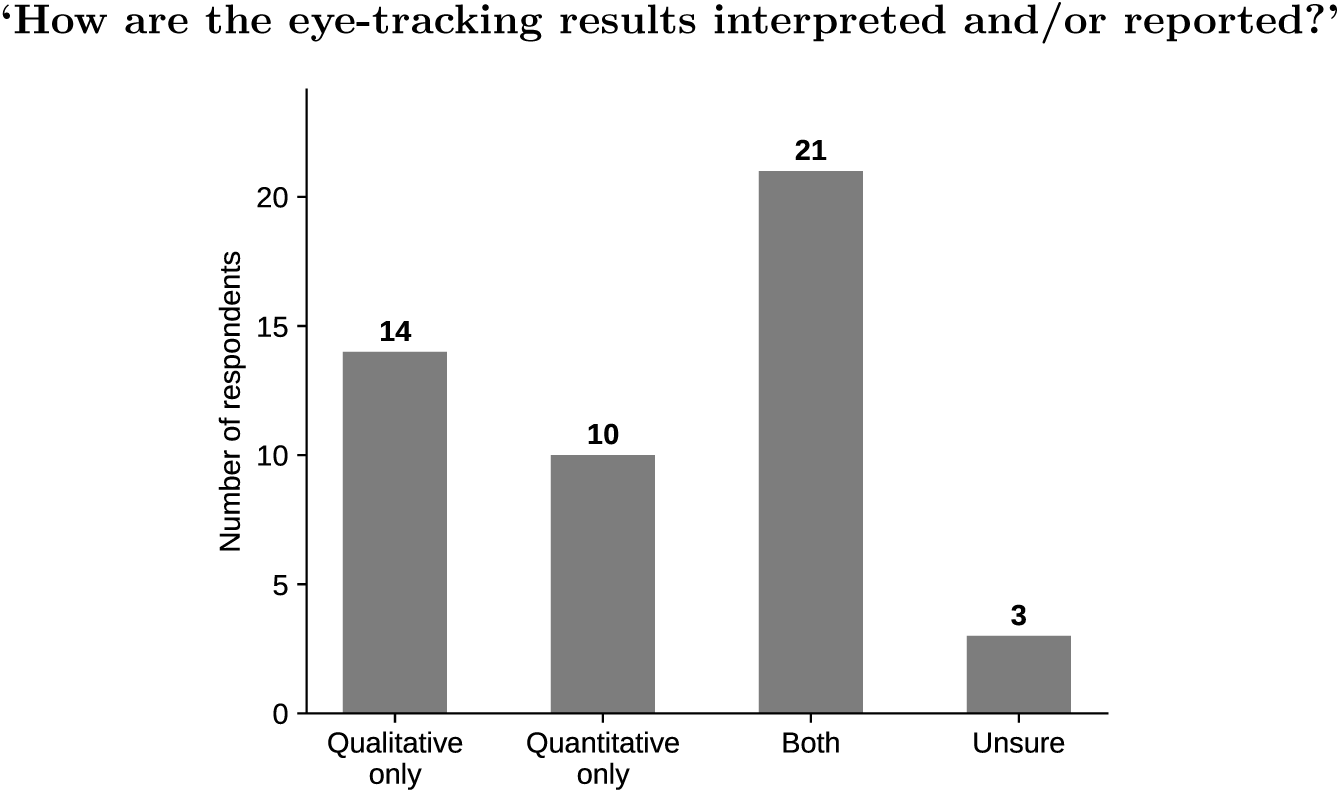
Type of interpretation and or reporting of eye-tracking results, from all respondents who answered this question (n=48) - including non-clinicians

**Fig. 5.**
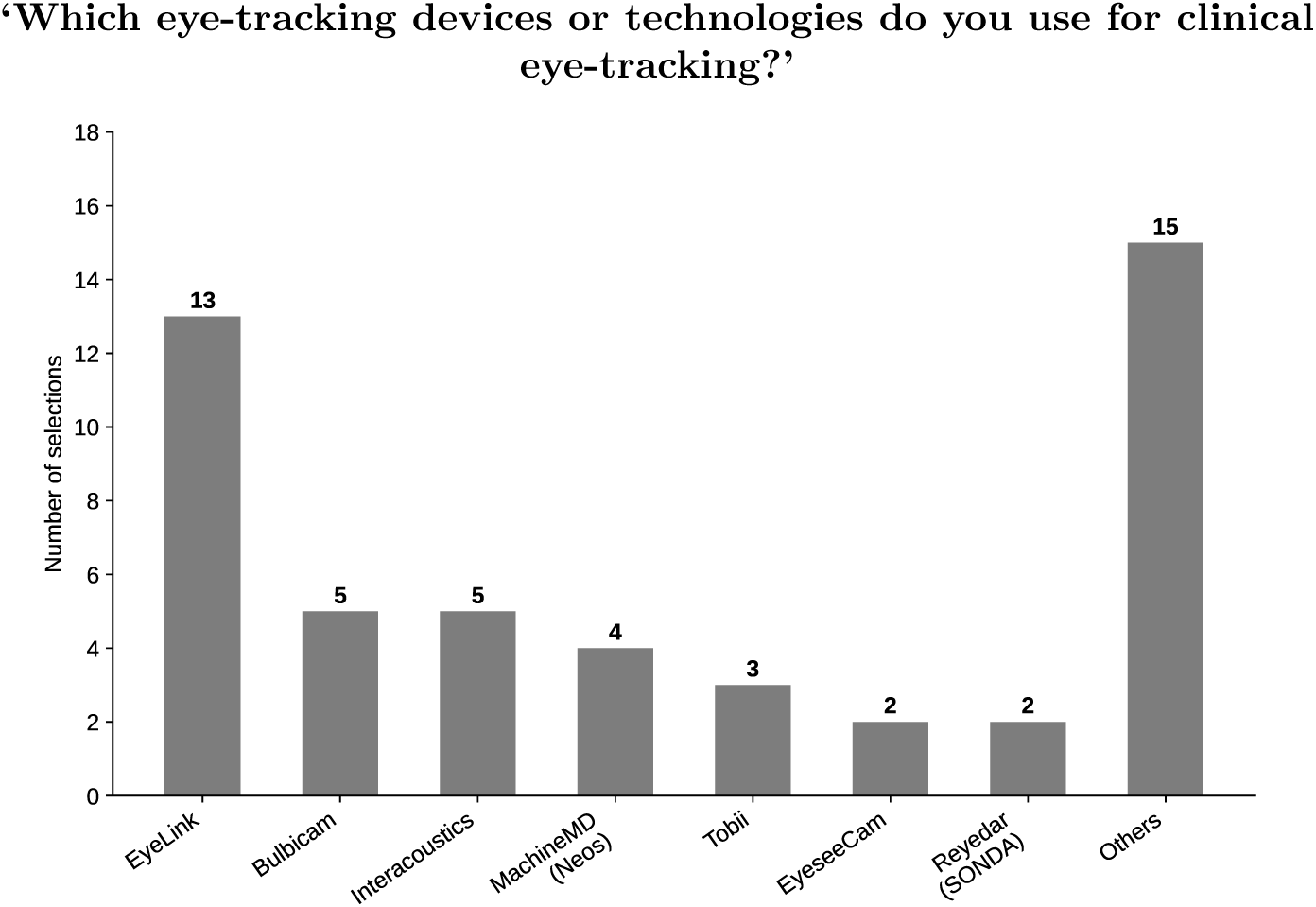
Eye-tracking devices used in clinical practice. Five respondents selected more than one device; four selected two devices, and one selected four devices. Devices selected only once are included in ‘Others’, together with free-text responses that could not be assigned to a named system

### Barriers to adoption

The most commonly identified barriers to clinical eye-tracking were eye-trackers being considered primarily research tools, considered unsuitable for performing the assessments, and no access to an eye-tracker, with limited time, cost, and lack of familiarity with eye-tracking also identified, see Figure 6.

**Fig. 6.**
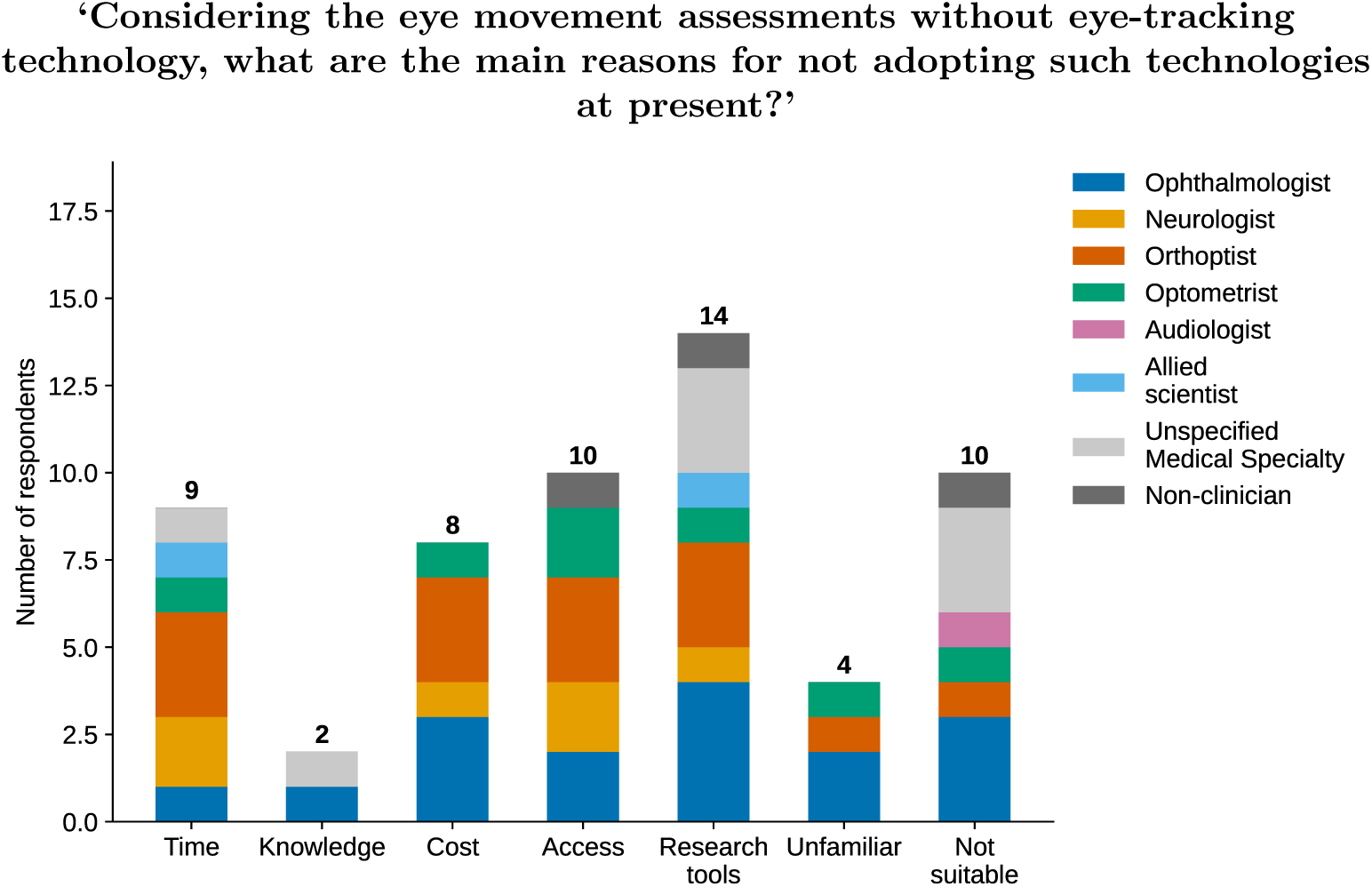
Barriers to the adoption of clinical eye-tracking, stratified by speciality. A respondent reporting both clinical science and optometry is counted here as an Optometrist. Respondents were asked to select any applicable barriers from a list of seven coded categories. Multiple responses were per-mitted; 14 selected more than one barrier. Five respondents selected ‘Other’ and are not represented in the plot: one Orthoptist noted having access to an eye-tracker but managing some cases without it; one Ophthalmologist described performing eye-tracking, rather than identifying a structural barrier; two Audiologists - one with no further detail, one describing situational device unavailability; and one non-clinician with no further detail

### Clinical guidelines

Awareness of guidelines for clinical eye-tracking was answered by 48 respondents. The majority of respondents were not aware of any existing guidelines for clinical eye-tracking (71%, 34/48), 21% (10/48) were aware of guidelines, with one (2%, 1/48) responding ‘yes, but we do not use them’ and 9 (19%, 9/48) responding ‘yes and we use them’. Four respondents (8%, 4/48) answered ‘other’, but gave insufficient information to include and categorise their responses as yes or no.

Of those that gave additional details of the existing guidelines they were aware of (free text), these were reported as from an institution (1/9), from a manufacturer (4/9), evidence-based (2/9), they cited documentation that was not relevant to clinical eye-tracking (4/9) or the guidelines were not stated (2/9). Some respondents reported multiple sources of existing guidelines. Existing guidelines were reported to range from ‘somewhat sufficient’ to ‘completely insufficient’.

The majority of respondents - 93% (43/46) - responded that guidelines for clinical eye-tracking should exist, and 7% (3/46) thought they should not. Reasons cited for the view that they should not exist can be summarised as: the complex and multi-factorial assessment of patients requires an individualised approach for each patient rather than a text book or prescribed approach; oculomotor assessments being different from electrophysiology (International Society for Clinical Electrophysiology of Vision, ISCEV); guidelines would be limiting; and it being a manufacturer role rather than a clinician role to develop guidelines. Responses from those that thought guidelines should exist included a wide range of suggested scenarios, conditions, or eye movements that such guidelines should cover, see Table 2.

**Table 2.** Thematic analysis of free-text responses to the question of which eye movement types, scenarios or conditions clinical eye-tracking guidelines should cover.

| Theme | Specific items mentioned | *n* |
| --- | --- | --- |
| Eye movement types | Saccades (pro-, anti-, double-step), smooth pursuit, nystagmus, fixation / fixation abnormalities, vergence, optokinetic nystagmus, vestibulo-ocular reflex, gaze holding, saccadic latency, central oculomotor signs, eye movement perimetry, pupillometry | 13 |
| Clinical conditions & patient populations | Parkinson’s disease / PSP / parkinsonism, MS, Alzheimer’s disease / neurodegenerative diseases, INO, myasthenia gravis, traumatic brain injury / stroke, nystagmus conditions, strabismus, gaze paresis, cranial nerve palsies, ataxia, metabolic / endocrine / neurological diseases, neuro-ophthalmic conditions, neuromuscular diseases, children, psychiatry, rehabilitation | 15 |
| Methodology & standardisation | Calibration, recording protocols, hardware / software setup and specifications, data export and analysis, reporting guidelines, inter-centre comparability, apparatus specifications, waveform interpretation, pre-testing procedures, data quality and error implications, minimal device requirements, consensus on visual stimuli | 16 |
| Clinical decisions | Thresholds for management decisions, when to repeat testing, normal variation vs. clinical significance, regulatory approval as primary outcome measure, patient expectations | 3 |
| Other / unclear | See note below | 7 |

*Note.* The seven responses coded as *Other / unclear* were: one describing electrophysiology; one referring to training clinical staff in monitoring attention and stress levels; and four responses too vague to code (‘diagnosis’; ‘research, post-op results’; ‘Yes for treatment and setting patient expectations’; and a response from a respondent who stated they were unfamiliar with clinical eye-tracking).

### Manufacturer responses

Nineteen respondents were from eye-tracker manufacturers (n=17) or people connected to the eye-tracking market (n=2). Of the eye-tracker manufacturers that responded regarding regulatory approval (n=16), 69% (11/16) reported that they market device(s) that have regulatory approval for clinical use and 44% (7/16) reported that they market device(s) without regulatory approval for clinical use, while two respondents reported marketing both approved and unapproved devices.

Of all those categorised as manufacturer responses, 68% (13/19) reported – to the best of their knowledge – how they believed analyses of eye-tracking results are performed by their users, with multiple responses possible. A total of 20 responses were recorded. Forty five percent of responses (9/20 responses) reported commercial software was used for analysis, 35% (7/20 responses) reported users own software was used, 10% (2/20 responses) reported open source software was used, 5% (1/20 responses) reported their users were not thought to be analysing the results and 5% (1/20 responses) were unsure how the analysis was performed.

### Professional body responses

Eight respondents indicated they were involved with a professional body or represented an organisation. Sixty-three percent (5/8) gave the name of their professional body, which were confirmed to be recognised organisations related to clinical practice; however names were withheld in accordance with the consent taken (see consent and survey questions in Online Resource 2). Three responded to the question “To the best of your knowledge, do your members have any concerns or issues specific to the use of eye-tracking technology clinically?”; one with ‘No’, one with ‘Yes’, and one with ‘Unsure’. In terms of which clinical specialisations their members represented, one responded with optometry, and one with ophthalmologists (with those performing eye-tracking being neuro-ophthalmologists). In response to the question “To the best of your knowledge, how are the eye-tracking results interpreted and or reported by your members?”, one responded quantitatively - by reporting numerical results, one reported both quantitatively and qualitatively - by observing the recorded information and describing the findings, and one reported that they were unsure how this is done.

## Discussion

This study aimed to characterise the current landscape of clinical eye-tracking through a survey of users and stakeholders. As far as we are aware, this is the first survey attempting to characterise the broad spectrum of clinical domains using eye-tracking.

### How are eye-trackers used clinically?

A diverse range of clinicians responded to the survey, including optometrists, ophthalmologists, orthoptists, neurologists, audiologists and allied scientists. Of those who responded, the majority assess eye movements on a daily basis, but use eye-tracking rarely (less than 10%) to moderately (10-60%) as part of their eye movement assessments. This is likely biased towards those with an interest in this area rather than faithfully representing the rates of eye-tracking usage across clinical professions as a whole. Nevertheless, it does demonstrate that there is scope for clinical eye-tracking across a broad range of use cases. A representative range of modern trackers were reported, with analysis undertaken by both commercial and bespoke software. There are no standardised clinical eye movement protocols with normative data. The adoption of calibration and validation protocols varies by region, use case, and clinical workflow. In practice, clinicians often have limited time or training to assess the quality of each recording or to adjust default settings for individual patients.

Although VOG has become the predominant automated method for clinical eye movement recording, other technologies retain selective value, for example, when torsional eye movement is relevant, as few VOG systems address this. EOG and its clinical derivative electronystagmography (ENG) are still used in specific contexts – particularly for caloric and rotational testing, or where patients cannot tolerate infrared cameras, as well as in sleep or other contexts where the eye is closed. EOG may also be the preferred method of eye-tracking in clinics that already perform electrodiagnostic tests, for example, when recording Visual Evoked Potentials from patients with suspected albinism. The technique is robust and low cost, but also limited by drift and lower spatial precision [10, 31]. Guidance documents such as those from the British Society of Audiology [11, 12] still reference EOG or ENG as acceptable alternatives to VOG under defined conditions. Magnetic SSC systems remain in use mainly within specialised centres requiring high temporal and spatial accuracy, where legacy research provides benchmarks [36, 64] or where ground-truth recordings are required for the development or validation of new clinical measures [34].

### Eye-tracking clinical guidelines

Technical capability has outpaced clinical standardisation. Although modern eye-tracking systems can measure fine-grained temporal dynamics across multiple degrees of freedom, there is considerable uncontrolled variance in algorithms for analysis between systems, and in data quality between systems and indeed between patients, while clinicians operate under constraints of time, training, and workflow. Many clinicians interpret eye-tracking outputs indirectly—through system-generated metrics or software classifications—without scope for routine inspection of raw traces. Others may continue to rely on visual observation supplemented by selective recording, especially in time-pressured environments. These differing practices reflect variation in both familiarity and available equipment and infrastructure, leading to inconsistencies in diagnostic criteria and thresholds.

The practical consequence is that clinical results cannot be directly compared across devices, datasets, patient visits, or sites without harmonised performance criteria. Eye-tracker manufacturers such as SR Research and Tobii now provide detailed calibration and validation protocols for their systems [68, 76], but these remain proprietary rather than standardised, and are designed primarily for research rather than clinical contexts.

The survey results demonstrate that there is a desire and need for consensus guidelines with 93% of respondents to that question indicating that such guidelines should exist. This is not surprising given the utility of eye-tracking demonstrated in the literature. The development of clinician-focused guidelines would allow standardisation for diagnosis, monitoring of a wide range of conditions, and provide a common framework when referring patients to or between clinical specialities. Most respondents (71%) were unaware of any guidelines for clinical eye-tracking, and where guidelines were identified, these were most often manufacturer documentation or material not specific to clinical eye-tracking. This is consistent with the published landscape.

Despite decades of progress in eye movement recording, professional guidance on how to record, validate, and interpret these data for diagnosis or to provide clinically useful information remains fragmented. Internationally, the Bárány Society has issued consensus definitions for vestibular disorders [2, 71–73, 84], establishing what should be diagnosed but not prescribing how recordings should be acquired or quantified. In the United States, the American Academy of Otolaryngology–Head and Neck Surgery Foundation (AAO-HNSF) guideline for benign paroxysmal positional vertigo [7] provides standardised manoeuvres and management recommendations but does not define eye-movement recording standards. The 2021 U.S. Department of Defense and Veterans Affairs (DoD/VA) Clinical Recommendations [78] similarly emphasise vestibular triage and rehabilitation pathways without specifying eye-movement or data-quality metrics.

National and regional bodies have contributed complementary procedural frameworks. The British Society of Audiology (BSA) has published recommended procedures for caloric and positional testing [11, 13] and for eye-movement recordings [12]. These guidelines have been influential procedural templates in many clinical contexts over the last decade: they describe calibration routines, fixation suppression, and reporting indices, explicitly recommending VNG over EOG where available. Existing publications predate the current generation of eye-tracking tools and guidelines require periodic review as the technology changes. Reliance on institutional procedures or manufacturer protocols often substitutes for up-to-date, formal standards. Resources such as the Interacoustics Academy’s “Caloric Test: Deep Dive” [35] provide detailed, recording-based instruction, but are designed for users of their particular hardware at a particular stage of technical development.

The most cited (n=14) barrier to clinical uptake of eye-tracking technologies was that eye-tracking was considered purely a research tool. The development of clinical guidelines may change this situation in three ways. Firstly, it would provide concrete, actionable guidance for busy clinicians without their having to integrate information from a vast research literature. Secondly, they would also provide benchmarks for manufacturers, for regulatory compliance, research and development, and advertising purposes. Thirdly, guidelines could provide a framework under which multicentre reference (‘normative’) data could be generated and referenced across research, clinical practice, and technical development.

Barriers to wider integration are practical as well as conceptual. Formal education in the technical and applied aspects of eye-tracking is rare outside research settings. Existing guidelines provide procedural structure for specific tests, but limited direction on quantitative interpretation or error handling. As a result, clinicians may depend on proprietary software defaults or manufacturer recommendations for calibration validity, eye-movement event detection (fixations, saccades, smooth pursuit, blinks), and artefact filtering or rejection, even though the literature demonstrates the critical role of individual characteristics and environments as well as hardware in contributing error to the signal. Pending standardisation, cross-platform and multi-site data aggregation remains unreliable, and the large-scale normative databases that could transform diagnostic accuracy and monitoring remain out of reach.

Of interest were the three respondents who indicated that eye-tracking guidelines should *not* exist. The comments accompanying these responses came from clinicians who value their autonomy and independence in judgement when assessing eye movements and using clinical eye-tracking, and perceived standardised guidelines as a threat to independent use and interpretation. The initial development of any guidelines should take this into consideration, providing benchmarks rather than prescriptive protocols which may be unwelcome.

As eye-tracking technologies are increasingly employed across domains such as ophthalmology, neurology, audiology, optometry, psychiatry, and orthoptics, we aim to address fragmented clinical implementation and lack of universally adopted guidelines or common principles to support clinical practice. The intention of this survey was to characterise the perspectives and practical needs of clinicians and allied stakeholders. The goal is to address these needs with consensus as the guiding principle for guideline development.

### Limitations

How accurately the number of respondents to the survey represent those using eye-tracking clinically is unknown. Despite attempts to reach a wide audience and capture clinical eye-tracker use internationally, it is acknowledged that the results of this survey may under-represent, or may closely represent, the numbers involved in practice. Several respondents (n=43) did not answer some or all of the survey questions, despite initially reporting they understood the participant information. It is unknown whether these were potentially relevant responses that were not captured, if these were non-clinicians or others who wanted to view but not answer questions.

This questionnaire required participants to select how they were involved in clinical eye-tracking under a fixed set of possible responses that aimed to cater to clinicians, researchers, manufacturers, and members of professional bodies. However, the lines between these roles are somewhat arbitrary and may not encompass mixed roles or roles that do not fall neatly into these categories. In many clinical settings, distinct categories of, e.g., research and clinical practice may not be reflective of everyday clinical work. Whether this may have influenced responses is unclear. Another issue seen retrospectively is that we asked for responses from those involved in clinical eye-tracking. This excluded clinicians who *aspire* to use eye-tracking in practice, but have not yet done so.

The survey was piloted to ensure questions were understandable, but it may be possible to improve some questions in the future based on the results presented. For example, respondents were asked if they ‘interpreted’ eye-tracking results; this may have been better divided into ‘process eye-tracking data’ and ‘use eye-tracking results to make clinical decisions’.

The present survey provides a useful characterisation of those currently involved in clinical eye-tracking and reports a majority wish for guidelines or recommendations to support eye-tracking in clinical practice. Repeating the survey in the future would allow an estimate of the growth of the field. Future versions of the survey should:

– Ask for the specific tests that are performed during clinical eye-tracking
– Ask for the amount of experience respondents have with clinical eye-tracking
– Consider whether there is value in distinguishing between respondents who are involved in vestibular neurology; audiology; and ear, nose, and throat specialities
– Consider whether there is value in distinguishing between respondents who are involved in ophthalmology; neurology; and neuro-ophthalmology specialities
– Consider including responses from those who would like to use eye-tracking clinically, but are unable to do so

## Conclusions

Overall, the present survey characterises a diverse set of users and stakeholders involved in clinical eye-tracking. Across a broad spectrum of application areas, there is wide support for the introduction of clinical guidelines for the use of eye-tracking technology from those using eye-trackers regularly, those using them occasionally, and those who perceive barriers to their clinical use. A small number of survey respondents do not support guideline development citing concerns about autonomy. For guidelines to be effective, it will be important to emphasise that guidelines do not override or replace the individual responsibility or judgement of clinicians in making appropriate decisions in the circumstances of their patients.

## Data Availability

All data produced in the present study are available upon reasonable request to the authors.

## Declarations

– **Funding**: This study was unfunded. The following authors have declared funding sources: R.S.M. is funded by Silmäsäätiä, Grant No. 20250022.
– **Competing interests**: M.J.D. is the General Coordinator of ISCET, recently joined the editorial board of *Documenta Ophthalmologica*. R.S.M. serves as the Vice President/Regional Coordinator for Europe & Africa Regions for ISCET. A.D. serves as the Vice President/Regional Coordinator for Australasia Regions for ISCET. S.C. is the Spokeperson for ISCET. F.S. is a paid consultant to Definium Therapeutics Inc. (formerly MindMed).
– **Ethical approval** for the study was granted by the University of Sheffield (UoS064587). All respondents gave consent to participate after reading the participant information and prior to completing any of the survey questions. Numbers of those accessing the survey but not fully consenting to all statements (and therefore not proceeding to the survey questions) are reported in the manuscript for transparency.
– **Consent for publication**: All authors gave explicit consent for the final version to be submitted, with their co-authorship.
– **Data availability**: G.A. has ethical approval for an anonymised data set to be uploaded to ORDA (Online Research Data repository, University of Sheffield).
– **Materials availability**: All materials relevant to this study have been presented as supplementary materials.
– **Code availability**: Plotting code available on request
– **Author contributions**: F.B.M. and G.A. contributed equally to the work. G.A. led the project, coordinated meetings and survey distribution, drafted the survey for review, wrote the methods, and co-authored results and discussion. F.B.M. conducted literature review, wrote introduction (comments from G.A., R.S.M., M.J.D., A.D., S.C.), drafted article, designed data handling, and coded data (together with D.G. and M.J.D., checked by S.C., G.A.), analysed and plotted data, co-authored results and discussion, prepared supplementary information documents. All co-authors commented on and edited the survey and the draft article.

## Supplementary information

***Online Resource 1 (ESM 1.pdf): list of organisations contacted*.**

***Online Resource 2 (ESM 2.pdf): survey questions, as administered*.**

***Online Resource 3 (ESM 3.pdf): data handling and inclusion criteria*.**

## Acknowledgements

Thank you to members of ISCEV and ISCET who contributed to the conceptualisation and interpretation of the survey, and to the survey working group members who supported its early development. We would like to acknowledge all those who shared, distributed and completed the survey.

## Notes

### Author Declarations

Ethical approval for the study was granted by the University of Sheffield (UoS064587). All respondents gave consent to participate after reading the participant information and prior to completing any of the survey questions. Numbers of those accessing the survey but not fully consenting to all statements (and therefore not proceeding to the survey questions) are reported in the manuscript for transparency.

