## Supplementary Information 1 for "A survey and review of eye-tracking in clinical practice"

Electronic Supplementary Material to:

*Documenta Ophthalmologica*

Fiona B.Mulvey,,
Onyekachukwu Mary-Anne Amiebenomo, Denize Atan, Siyuan Chen,
Amanda Douglass, Matt J. Dunn, Daniel Goldstone, Rasha Sameer Moustafa, Frederic Shic,
Gemma Arblaster*,

*Corresponding author

---

Organisations identified as relevant stakeholders and contacted with an invitation to complete the survey, grouped by category (n=106).

### Eye movement disorders

| Organisation |
|---|
| NUKE - Nystagmus UK Eye Research Group (within Nystagmus Network) (UK) |
| ANN - American Nystagmus Network (USA) |

### Orthoptics

| Organisation |
|---|
| IOA - International Orthoptic Association (International) |
| OCE - Orthoptistes de la Communauté Européenne (Europe) |
| BIOS - British and Irish Orthoptic Society (UK) |

### Optometry

| Organisation |
|---|
| WCO - World Council of Optometry (International) |
| ECOO - The European Council of Optometry and Optics (Europe) |
| AFCO - The African Council of Optometry (Africa) |

| |
|---|
| GOC - General Optical Council (UK) |
| The College of Optometrists (UK) |
| Association of Optometrists (UK) |
| AAO - The American Academy of Optometry (USA) |
| Optometry Australia (AUS) |

### Ophthalmology

| **Organisation** |
|---|
| ICO - International Council of Ophthalmology (International) |
| ARVO - The Association for Research in Vision and Ophthalmology (USA) |
| American Academy of Ophthalmology (USA) |
| The Royal College of Ophthalmologists (UK) |
| RANZCO - The Royal Australian and New Zealand college of Ophthalmology (AUS) |
| AAPOS - American Association for Pediatric Ophthalmology and Strabismus (USA) |
| BIPOSA - British and Irish Paediatric Ophthalmology and Strabismus Association (UK) |

### Neuro-ophthalmology

| **Organisation** |
|---|
| EUNOS - European Neuro-Ophthalmology Society (Europe) |
| UK Neuro-Ophthalmology Special Interest Group (UK) |
| UKNOS - UK Neuro-Ophthalmology Society (UK) |
| British and Irish Neuro-Ophthalmology Club (UK) |
| NANOS - North American Neuro-Ophthalmology Society (USA) |
| NOSA - The Neuro-Ophthalmology Society of Australia (AUS) |

### Neurology and neuroscience

| **Organisation** |
|---|
| WFN - World Federation of Neurology (International) |
| EBC - European Brain Council (Europe) |
| SFN - Society for Neuroscience (USA) |

#### Neurodiversity and reading disability

| Organisation |
| --- |
| INA - International Neurodiversity Alliance (International) |
| ION - Institute of Neurodiversity (International) |
| COVD - College of Optometrists in Vision and Development (USA) |
| FNIH Biomarkers consortium [Autism Biomarkers Consortium for Clinical Trials (ABC-CT)] (USA) |
| International Society for Autism Research (INSAR) (INternational) |
| Autism Science Foundation (ASF) (USA) |

#### Neuropsychology

| Organisation |
| --- |
| INS - International Neuropsychological Society (International) |
| The British Psychological Society (UK) |

#### Psychiatric conditions and mental health

| Organisation |
| --- |
| WFMH - World Federation of Mental Health (International) |

#### Visual electrophysiology

| Organisation |
| --- |
| ISCEV - International Society for Clinical Electrophysiology of Vision (International) |
| BriSCEV - British Society for Clinical Electrophysiology of Vision (UK) |

#### Dizziness (hearing and balance)

| Organisation |
| --- |
| The Barany Society - The International Society of Neuro-otology (International) |

| |
|---|
| ARO - Association for Research in Otolaryngology (USA) |
| AAO-HNS - American Academy of Otolaryngology, Head and Neck Surgery (USA) |
| IFOS - International Federation of Otorhinolaryngological Societies (International) |
| BSO - British Society of Otology (UK) |

### Audiology / ENT

| Organisation |
|---|
| ISA - International Society of Audiology (International) |
| BAA - British Academy of Audiology (UK) |
| BSA - British Society of Audiology (UK) |

### Concussion

| Organisation |
|---|
| Concussion.org - International Concussion Society |
| Concussion in Sport Group (CISG) |

### Vision (general)

| Organisation |
|---|
| VSS - Vision Science Society (USA) |
| Eye-tracking User Group (UK) |
| SIGCHI - Special Interest Group on Computer-Human Interaction (international) |
| BOMG - British oculomotor Group (UK) |

### Eye-tracker manufacturers: clinical

| Organisation |
|---|
| NATUS (ICS Impulse) |
| Interacoustics (EyeSeeCam / VisualEyes) |
| Neurofit (NeuroFit One) |
| Bulbitech (BulbiCAM) |
| Inventis (Synapsys/Nystalyze) |

| |
|---|
| Suricog [E(ye)BRAIN] |

### Eye-tracker manufacturers: non-clinical

| **Organisation** |
|---|
| SR Research Ltd. (EyeLink) |
| Tobii (Tobii Pro) |
| Cambridge Research Systems |
| FOVE |
| Pupil Labs (Neon) |
| Gazepoint (Gazepoint GP3) |
| Smart Eye / iMotions (SmartEye AI-X, SMI) |
| EyeSeeTec (EyeSeeCam) |
| EyeGaze (EyeGaze Edge Link) |
| Eyetech |
| Positive Science |
| Plusoptix |
| Seeing Machines |
| Arrington Research (ViewPoint Eye Tracker systems) |
| ASL - Applied Science Laboratories (EYE-TRAC) |
| Reality Labs |
| Eye Square |
| View Point System |
| RealEye (Webcam Eye-Tracking) |
| eyeLOGIC |
| VPixx Vision Science Solution (TRACKPixx) |
| SeeTrue Technologies |

### Eye-tracker manufacturers: software-based

| **Organisation** |
|---|
| Visage Technologies AB |

### Consumer-electronics manufacturers

| **Organisation** |
|---|
| Meta |
| Varjo |
| Pimax |

### Regulatory bodies

| Organisation |
|---|
| FDA - Food and Drug Administration (USA) |
| MHRA - Medicines and Healthcare Products Regulatory Agencies (UK) |
| TGA - Therapeutic Goods Administration (AUS) |

### Funding agencies: neuroscience

| Organisation |
|---|
| NIMH - National Institute of Mental Health (USA) |
| NINDS - National Institute of Neurological disorders and Stroke (USA) |
| American Brain Foundation (INA Research Fund) |

### Funding agencies: vision research

| Organisation |
|---|
| NEI - National Eye Institute (USA) |

### Funding agencies: ENT

| Organisation |
|---|
| NIDCD - The National Institute on Deafness and other Communication Disorders (USA) |

### Funding agencies: other

| Organisation |
|---|
| UKRI - UK Research and Innovation |
| NIH - National Institute of Health (USA) |
| Fight for Sight (UK) |
| Nystagmus Network (UK) |

### Additional groups

| Organisation |
| --- |
| ISCET - International Society for Clinical Eye Tracking |
| EYE-MOVEMENT list on JISCmail |
| Eye tracking experts and researchers at INT group, UEF, Finland |
| ETRA |
| Blickshift GmbH |
| Smart Eye AB |
| DM executive members: ISLRR: International Society for Low vision Rehabilitation and research: |
| Arabic speaking Ophthalmologist Group |
