## Supplementary Information 2 for "A survey and review of eye-tracking in clinical practice"

Electronic Supplementary Material to:

*Documenta Ophthalmologica*

Fiona B.Mulvey,,
Onyekachukwu Mary-Anne Amiebenomo, Denize Atan, Siyuan Chen, Amanda Douglass, Matt J. Dunn, Daniel Goldstone, Rasha Sameer Moustafa, Frederic Shic, Gemma Arblaster*,

*Corresponding author

---

This document reproduces the survey instrument as administered. The survey was implemented in Qualtrics (Qualtrics, Provo, UT, USA). Items annotated *Shown if* were displayed conditionally on the respondent's earlier answers. Response-capture types are given in brackets. Respondents answering as manufacturers were shown the manufacturer questions before the questions asked of all respondents.

### Consent statements

Please read the following statements and tick the box if you agree with each statement.
**Taking part in the research: How are eye-trackers used clinically?**

- I have read and understood the participant information (version 2).
- I understand I can contact the researcher by email if I have questions about the survey.
- I agree to take part in the survey and understand that taking part will involve me completing a survey either anonymously (as an individual) OR by sharing the name of the organisation I am completing the survey on behalf of (for example manufacturer or professional body).
- I understand that taking part in the survey is voluntary and I cannot withdraw my anonymous responses, if I am an individual completing the survey, but I can withdraw my responses prior to anonymisation, if I am completing the survey on behalf of an organisation (e.g. professional body or manufacturer). I do not have to give a reason for withdrawing and there will be no adverse consequences if I choose to withdraw my responses.

**How my information will be used during and after the survey:**

- I understand that any survey responses containing organisation names will not be shown to people outside the ISCET survey working group.
- I understand and agree that my anonymous responses and my words may be quoted anonymously in publications, reports, web pages and other research outputs.

**So that the information you provide can be used legally by the researchers:**

- I agree to assign the copyright I hold in any materials generated as part of this project to The University of Sheffield.

### Section A. Screening

1. Which of the following best describes your involvement with clinical eye tracking? *[single choice]*

*If more than one option is applicable to you, please select the option that best describes your involvement and complete the survey in that capacity. You may complete the survey multiple times in alternative capacities (if relevant), by selecting an alternative option.*

- I perform clinical eye tracking and interpret the results
- I perform clinical eye tracking but do not interpret the results
- I do not perform clinical eye tracking, but interpret or use the results recorded by others
- I am involved with a professional body
  - Name of professional body *[free text; may be left blank]*
- I am involved with a manufacturer of devices or technology related to clinical eye tracking
  - Name of manufacturer / company *[free text; may be left blank]*
- Other *[free text]*

### Section B. Clinician questions

1. Who would interpret the eye tracking results? *[single choice]*

   *Shown if: "I perform clinical eye tracking but do not interpret the results" selected at Q1.*

   - Medically trained doctor / Ophthalmologist
   - Other vision specialist (Clinical Scientist or other qualified person in field of vision science)
   - Orthoptist
   - Optometrist
   - Eye tracking technology specialist
   - Computer science expert
   - Other *[free text]*

2. Which of the following do they use? *[multiple choice]*

   *Shown if: "I do not perform clinical eye tracking, but interpret or use the results recorded by others" selected at Q1.*

   - Eye tracking devices or technology with regulatory approvals for clinical use
     - Name of devices or technologies *[free text]*
   - Eye tracking devices or technology without regulatory approval for clinical use, but has been applied by others to clinical eye tracking
     - Name of devices or technologies *[free text]*
   - Other *[free text]*

3. Which of the following do you use? *[multiple choice]*

   *Shown if: "I perform clinical eye tracking and interpret the results"* or *"I perform clinical eye tracking but do not interpret the results" selected at Q1.*

   - Eye tracking devices or technology with regulatory approvals for clinical use
     - Name of devices or technologies *[free text]*
   - Eye tracking devices or technology without regulatory approval for clinical use, but has been applied by others to clinical eye tracking

    - Name of devices or technologies *[free text]*
- Other *[free text]*

4. Which of the following best describes your professional role? *[multiple choice]*
    - I am clinically qualified
    - I am clinically registered
    - Other *[free text]*

5. What is your clinical discipline? *[multiple choice]*
    - Medically trained doctor
        - Medical speciality *[free text]*
    - Clinical scientist
    - Orthoptist
    - Other Allied Health Professional (AHP) *[free text]*
    - Optometrist
    - Other vision specialist *[free text]*
    - Other clinician *[free text]*

6. Approximately how often do you perform eye movement assessments in the course of your clinical duties? *[free text]*

7. Of the times you perform eye movement assessments, approximately what proportion of these use eye tracking technology? *[free text]*

8. Considering the eye movement assessments without eye tracking technology, what are the main reasons for not adopting such technologies at present? *[multiple choice]*
    - Unfamiliar with eye tracking
    - No access to an eye tracker
    - Would like to perform eye tracking — but they are too expensive
    - Access to an eye tracker — but have limited time to perform eye tracking
    - Access to an eye tracker — but limited knowledge of how to use eye tracking equipment
    - The information generated by an eye tracker would not be used clinically
    - Eye trackers are predominantly research tools
    - Eye trackers are not safe to use clinically
    - To my knowledge, there is currently no suitable eye tracker to perform our assessment
    - Other *[free text]*

9. Do you have any concerns or issues specific to the use of eye tracking technology clinically? *[single choice, plus free text]*
    - Yes
    - No
    - Unsure
    - Additional comments *[free text]*

10. How are the eye tracking results interpreted and/or reported (by you or by others)? *[multiple choice]*
    - Qualitatively — by observing the recorded information and describing the findings
    - Quantitatively — by reporting numerical results
    - Other
    - I am unsure how this is done

11. How are the eye tracking results analysed? *[multiple choice]*
    - Commercial software
    - Own software — produced in house
    - Using open source code
    - No analysis is performed
    - I am unsure how the analysis is done
    - Other *[free text]*

### Section C. Manufacturer questions

*Shown if: "I am involved with a manufacturer of devices or technology related to clinical eye tracking" selected at Q1. These items were presented before the questions asked of all respondents (Section E).*

1. Do you produce or market any of the following? *[multiple choice]*
    - Eye tracking devices or technology with regulatory approvals for clinical use
        - Name of devices or technologies *[free text]*
    - Eye tracking devices or technology without regulatory approval for clinical use, but has been applied by others to clinical eye tracking
        - Name of devices or technologies *[free text]*
    - Other *[free text]*

2. Do you provide any guidelines for clinical eye tracking to your users? *[single choice, with follow-ups]*
    - Yes
        - What are these guidelines for? *[free text]*
        - If these are published online, please provide a link *[free text]*
        - If no link is available, please provide a brief description and the source of the guideline *[free text]*
    - No
        - Reasons for this *[free text]*
    - Other *[free text]*

3. To the best of your knowledge, do your users have any concerns or issues specific to the use of eye tracking technology clinically? *[single choice, plus free text]*
    - Yes
    - No

- Unsure
- Additional comments *[free text]*

4. To the best of your knowledge, how are the eye tracking results interpreted and/or reported by your users? *[multiple choice]*
   - Qualitatively — by observing the recorded information and describing the findings
   - Quantitatively — by reporting numerical results
   - Other
   - I am unsure how this is done

5. To the best of your knowledge, how are the eye tracking results analysed by your users? *[multiple choice]*
   - Commercial software
   - Own software — produced in house
   - Using open source code
   - No analysis is performed
   - I am unsure how the analysis is done
   - Other *[free text]*

#### Section D. Professional body questions

*Shown if: "I am involved with a professional body" selected at Q1.*

1. What are the clinical discipline(s) of your members? *[free text]*

2. How many members do you represent? *[free text]*

3. Approximately what proportion of your members perform eye movement assessments in the course of their clinical duties? *[free text]*

4. Of your members performing eye movement assessments, approximately what proportion of these use eye tracking technology? *[free text]*

5. What are the clinical discipline(s) of your members that perform eye tracking? *[free text]*

6. To the best of your knowledge, which of the following do your members use? *[multiple choice]*
   - Eye tracking devices or technology with regulatory approvals for clinical use
     - Name of devices or technologies *[free text]*
   - Eye tracking devices or technology without regulatory approval for clinical use, but has been applied by others to clinical eye tracking
     - Name of devices or technologies *[free text]*
   - Other *[free text]*

7. Considering your members who perform eye movement assessments without eye tracking technology, what are the main reasons for not adopting such technologies at present? *[multiple choice]*

- Unfamiliar with eye tracking
- No access to an eye tracker
- Would like to perform eye tracking — but they are too expensive
- Access to an eye tracker — but have limited time to perform eye tracking
- Access to an eye tracker — but limited knowledge of how to use eye tracking equipment
- The information generated by an eye tracker would not be used clinically
- Eye trackers are predominantly research tools
- Eye trackers are not safe to use clinically
- To my knowledge, there is currently no suitable eye tracker to perform our assessment
- Other *[free text]*

8. To the best of your knowledge, do your members have any concerns or issues specific to the use of eye tracking technology clinically? *[single choice, plus free text]*
   - Yes
   - No
   - Unsure
   - Additional comments *[free text]*

9. To the best of your knowledge, how are the eye tracking results interpreted and/or reported by your members? *[multiple choice]*
   - Qualitatively — by observing the recorded information and describing the findings
   - Quantitatively — by reporting numerical results
   - Other
   - I am unsure how this is done

10. To the best of your knowledge, how are the eye tracking results analysed by your members? *[multiple choice]*
    - Commercial software
    - Own software — produced in house
    - Using open source code
    - No analysis is performed
    - I am unsure how the analysis is done
    - Other *[free text]*

#### Section E. "Other" respondents

*Shown if: "Other" selected at Q1.*

1. Which of the following do you use? *[multiple choice]*
   - Eye tracking devices or technology with regulatory approvals for clinical use
     - Name of devices or technologies *[free text]*
   - Eye tracking devices or technology without regulatory approval for clinical use, but has been applied by others to clinical eye tracking

  - Name of devices or technologies *[free text]*
- Other *[free text]*

2. How would you describe your professional role? *[free text]*

3. Approximately how often do you perform eye movement assessments in the course of your role? *[free text]*

4. Of the times you perform eye movement assessments, approximately what proportion of these use eye tracking technology? *[free text]*

5. Considering the eye movement assessments without eye tracking technology, what are the main reasons for not adopting such technologies at present? *[multiple choice]*
   - Unfamiliar with eye tracking
   - No access to an eye tracker
   - Would like to perform eye tracking — but they are too expensive
   - Access to an eye tracker — but have limited time to perform eye tracking
   - Access to an eye tracker — but limited knowledge of how to use eye tracking equipment
   - The information generated by an eye tracker would not be used clinically
   - Eye trackers are predominantly research tools
   - Eye trackers are not safe to use clinically
   - To my knowledge, there is currently no suitable eye tracker to perform our assessment
   - Other *[free text]*

6. Do you have any concerns or issues specific to the use of eye tracking technology clinically? *[single choice, plus free text]*
   - Yes
   - No
   - Unsure
   - Additional comments *[free text]*

7. How are the eye tracking results interpreted and/or reported (by you or by others)? *[multiple choice]*
   - Qualitatively — by observing the recorded information and describing the findings
   - Quantitatively — by reporting numerical results
   - Other
   - I am unsure how this is done

8. How are the eye tracking results analysed? *[multiple choice]*
   - Commercial software
   - Own software — produced in house
   - Using open source code
   - No analysis is performed
   - I am unsure how the analysis is done
   - Other *[free text]*

### Section F. Questions to all respondents

1. Are you aware of any guidelines for clinical eye tracking currently? *[single choice, with follow-ups]*

   *If you are completing the questionnaire as a manufacturer, please answer this question considering "other guidelines" (i.e. those not provided by your company/organisation).*

   - Yes — but we do not use them
     - What are these guidelines for? *[free text]*
   - Yes — and we use them
     - What are these guidelines for? *[free text]*
   - No
   - Other *[free text]*

2. Please provide a link to any guidelines available. If no link is available, please provide a brief description and the source of the guideline. *[free text]*

   *Shown if: "Yes" answered at Q36.*

3. In your opinion, to what extent are current guidelines for clinical eye tracking sufficient? *[5-point scale]*

   - 1 — completely insufficient
   - 2 — somewhat insufficient
   - 3 — neither insufficient nor sufficient
   - 4 — somewhat sufficient
   - 5 — completely sufficient

4. Please give further comments about current clinical eye tracking guidelines. *[free text]*

5. Where they do not currently exist, do you think there is a need for guidelines for clinical eye tracking? *[single choice, with follow-ups]*

   - Yes
     - What scenarios, conditions or eye movements do you think there should be guidelines for clinical eye tracking? *[free text]*
     - What would you like guidelines for clinical eye tracking to contain? (e.g. specific test protocols or analysis procedures, information on suitable populations, and guidance to suitable eye tracking techniques, other) *[free text]*
   - No
     - Why? *[free text]*
   - Other *[free text]*

6. Is there anything else you would like to add which may be relevant to clinical eye tracking? *[free text]*

---

*This is the end of the survey.*
