## Supplementary Information 3 for "A survey and review of eye-tracking in clinical practice"

Electronic Supplementary Material to:

*Documenta Ophthalmologica*

Fiona B.Mulvey,,
Onyekachukwu Mary-Anne Amiebenomo, Denize Atan, Siyuan Chen, Amanda Douglass,
Matt J. Dunn, Daniel Goldstone, Rasha Sameer Moustafa, Frederic Shic,
Gemma Arblaster*,

*Corresponding author

---

This document describes how the survey responses were retrieved, cleaned, grouped, coded and analysed. Question numbers refer to the survey instrument as administered in Qualtrics (Online Resource 2).

### 1 Data retrieval

Survey results were downloaded from Qualtrics into a Microsoft Excel spreadsheet, which was uploaded to Google Drive and worked on as a Google Sheet. Respondent IP addresses were compared in order to identify possible duplicate responses.

### 2 Data cleaning

Data were inspected manually and processed according to the following steps.

1. Identifiable data provided by Qualtrics (IP addresses, and latitude and longitude) were removed.

2. Responses collected in survey preview mode — that is, during survey testing and prior to distribution — were removed.

3. Responses were then separated into the following groups:

   (a) Respondents who indicated that they had read and understood the participant information, responded to all of the statements relating to taking part in the research and to how their information would be used, and answered one or more questions in the survey.

   (b) Respondents who indicated that they had read and understood the participant information, responded to all of the statements relating to taking part in the research and to how their information would be used, but answered none of the survey questions.

   (c) Respondents who indicated that they had read and understood the participant information, responded to some but not all of the statements relating to taking part in the research and to how their information would be used, and therefore answered none of the survey questions (as they did not progress to the survey questions).

   (d) Respondents who indicated that they had read and understood the participant information, but answered none of the statements relating to taking part in the research and to how their information would be used, and therefore answered none of the survey questions (as they did not progress to the survey questions).

4. Responses (a) were analysed. Responses (b), (c) and (d) were counted and reported, but no results were available for analysis.

### 3 Grouping of responses

Responses were collated according to the answer given to Q4, “Which of the following best describes your involvement with clinical eye tracking?”

Because the survey branched on the answer to Q4, equivalent questions were presented to different respondent groups at different points in the instrument, and therefore appear as separate columns in the exported data. For example, Q7, Q15 and Q25 all ask which eye-tracking devices are used and whether those devices hold regulatory approval. Such columns were identified and collated. This process was repeated for each set of equivalent questions, giving the groupings below.

| **Content of question** | **Questions collated** |
|---|---|
| Devices used and regulatory approval | Q7, Q15, Q25, Q31, Q36 |
| Clinical speciality | Q8, Q23 |
| Frequency of eye-movement assessment | Q9, Q39 |
| Proportion of patients or members | Q10, Q29, Q40 |
| Barriers to adoption | Q11, Q32, Q41 |
| Concerns or issues | Q12, Q18, Q33, Q42 |
| Interpretation and reporting of results | Q13, Q19, Q34, Q43 |
| Analysis of results | Q14, Q20, Q35, Q44 |

### 4 Coding

Once the columns had been organised, the questions were examined for stratifications to which responses could be coded. Some stratifications follow directly from the multiple-choice options offered; others were derived by semantic analysis of the free-text answers provided. For example, Q51 asked which scenarios might require guidance to be developed; themes that appeared more than once (“saccades”, “pursuit”, “nystagmus”, “neurological disorders”, “palsies” and “strabismus”) were assigned columns so that they could be counted.

After the categories had been decided, the three-person team reviewing the data met to discuss any issues with the chosen categories and agreed any changes.

Stratifications were then coded with a 1 in each applicable column. This scheme was adopted so that a single response could belong to more than one category, or a single respondent could have more than one role or specialisation, and to facilitate plotting of the data. The only exception to this was with regards to calculating response rates as a percentage of respondents. In such cases, to avoid counting the same person twice, one specialisation was selected from those applicable.

The three-person data team subsequently reviewed the data and independently checked whether they agreed with the coding and with any discrepancies in the data. There were cases where free text responses contained an answer to a different question - such as, free text indicated that the respondent was a manufacturer, but they had coded themselves as a professional body - which was not the categorisation intended for ’professional body’. Any disagreements that could not be resolved within the data team were referred to the wider ISCET survey working group for consensus handling.

### 5 Analysis and plotting

Following agreement on the coding, the final spreadsheet was converted to plain text (`.csv`) to prevent artefacts arising across operating systems, machines and software. The `.csv` file, including all manual coding, was imported into Python for plotting, and plots were produced using a Python script (written in IDLE).

### 6 Additional notes on recoding

Three response-level recoding decisions were made:

- Where a respondent believed that the device they use was approved for clinical use, but named a device that is not in fact approved for clinical use, the response for "approved for clinical use" was changed to "no". This applied to one response.
- For Q8, one individual gave their speciality as "ENT"; this respondent was assigned to the "Neurologist" column.
- For Q8, two individuals gave their speciality as "Neuro-ophthalmologist" or "Neuro-ophthalmology"; both neurology and ophthalmology were coded for these respondents where multiple responses were possible. For cases where the goal was to calculate percentages of respondents, these respondents were assigned to the "Ophthalmologist" column to avoid being counted twice. They arguably fit in either category - so this decision was arbitrary and purely to avoid inflated respondent denominators in percentage response calculations.
